# Combined cerebrospinal fluid metabolomic and cytokine profiling in tuberculosis meningitis reveals robust and prolonged changes in immunometabolic networks

**DOI:** 10.1101/2023.06.26.23291676

**Authors:** Jeff Tomalka, Ashish Sharma, Alison G.C. Smith, Teona Avaliani, Mariam Gujabidze, Tinatin Bakuradze, Shorena Sabanadze, Dean P. Jones, Zaza Avaliani, Maia Kipiani, Russell R. Kempker, Jeffrey M. Collins

**Author notes:** Correspondence: Name: Jeffrey Tomalka, PhD; Jeffrey Collins, MD.

## Abstract

Much of the high mortality in tuberculosis meningitis (TBM) is attributable to excessive inflammation, making it imperative to identify targets for host-directed therapies that reduce pathologic inflammation and mortality. In this study, we investigate how cytokines and metabolites in the cerebral spinal fluid (CSF) associate with TBM at diagnosis and during TBM treatment. At diagnosis, TBM patients demonstrate significant increases versus controls of cytokines and chemokines that promote inflammation and cell migration including IL-17A, IL-2, TNFα, IFNγ, and IL-1β. Inflammatory immune signaling was strongly correlated with immunomodulatory metabolites including kynurenine, lactic acid, carnitine, tryptophan, and itaconate. Inflammatory immunometabolic networks were only partially reversed with two months of effective TBM treatment and remained significantly different versus control CSF. Together, these data highlight a critical role for host metabolism in regulating the inflammatory response to TBM and indicate the timeline for restoration of immune homeostasis in the CSF is prolonged.

## INTRODUCTION

Tuberculosis meningitis (TBM) is the most devastating manifestation of TB disease with high rates of mortality and neurological morbidity.(1) While the etiology of poor clinical outcomes in persons with TBM is multifactorial, including variable penetration of anti-TB drugs into brain and cerebrospinal fluid (CSF),(2) an aberrant immune response in the central nervous system (CNS) is thought to be a significant contributor to morbidity and mortality.(3) This is supported by early animal models of TBM, which revealed that severe immunopathology resulted from both live and dead bacilli.(4) Further, immune reconstitution in immunosuppressed hosts with TBM, such as those with HIV infection, is a risk factor for poor clinical outcomes.(5, 6) Corticosteroids, a therapy broadly targeting the immune dysregulation in TBM, remain the only adjunctive treatment found to reduce mortality and utilized in TBM management (7); however, they are only beneficial in a subset of patients.(8) A better understanding of the host immune response in TBM is critical to develop a new generation of immunomodulatory therapies that can reduce immunopathology and improve clinical outcomes.

As with other forms of meningitis, TBM is characterized by broad upregulation of proinflammatory cytokines in the CNS.(9) Yet the specific immune signaling cascades that lead to poor host outcomes remain poorly understood. While increased CSF concentrations of proinflammatory cytokines IL-1β, TNFα, IL-6, and IFNγ are associated with greater disease severity at diagnosis, studies to date have yielded conflicting results about their impact on long-term morbidity and mortality.(8, 10, 11) Some of this variability may be due to differences in host genetic (12) and metabolic factors (13, 14), which have generally gone unmeasured in studies of TBM. Indeed, differences in tryptophan and eicosanoid metabolism are the only host response pathways shown to impact TBM mortality.(8, 12–14) However, the relationship between metabolic and cytokine signaling in the CNS of TBM patients and the trajectory of these signaling networks during treatment remain largely unexplored.

Recent advances in metabolomics and high-density cytokine profiling now allow for high-resolution immunometabolic profiling of the CSF. Using an integrated approach with these two high-resolution methods, we sought to advance our understanding of the relationship between metabolism and cytokine signaling networks in the CNS of persons with TBM. We first sought to understand what metabolites and cytokines in the CSF are associated with TBM and TBM severity. We further aimed to determine how the host metabolic milieu in the CNS may modulate immune signaling by exploring the relationship between cytokines and metabolites in the CSF. Finally, we wanted to determine the time course for the resolution of these immunometabolic signaling cascades over the course of TBM treatment. Our overall goal was to enhance the understanding of host immunometabolic response networks to *Mycobacterium tuberculosis (Mtb)* invasion into the CNS and to elucidate their role in pathologic inflammation.(15)

## MATERIAL AND METHODS

### Setting and Participants with TBM

Persons with TBM were enrolled from the National Center for Tuberculosis and Lung Diseases (NCTLD) in Tbilisi, Georgia as part of a clinical pharmacology study evaluating the penetration of anti-TB drugs into the CNS.(2) Patients aged ≥16 years treated in the NCTLD adult TBM ward from January 2018 to December 2019 were eligible for inclusion. All patients suspected of having TBM underwent a lumbar puncture; acid-fast bacilli (AFB) staining, liquid and solid culture, and Xpert MTB/RIF assay were performed on CSF. CSF samples were taken at the time of TBM diagnosis and over the first 2 months of treatment. Treatment regimens were individualized based on treatment history, comorbidities, and drug susceptibility results when available.(16) All patients also received a 6–8-week course of dexamethasone (400–1200mg). Written informed consent was obtained from all study participants and study was approval was obtained from the institutional review boards of Emory University and the NCTLD.

### Asymptomatic Control Participants

De-identified CSF samples from asymptomatic persons enrolled as part of the Emory Healthy Brain Study were used as a control group for comparison to persons with TBM at diagnosis.(17) All study participants had previously provided written informed consent for use of their CSF samples in medical research.

### Metabolomics analysis

De-identified CSF samples were randomized by a computer-generated list into blocks of 40 samples prior to transfer to the analytical laboratory where personnel were blinded to clinical and demographic data. Thawed CSF (65 μL) was treated with 130 μl acetonitrile (2:1, v/v) containing an internal isotopic standard mixture (3.5 μL/sample), as previously described.(18) Samples were centrifuged and supernatants were analyzed using an Orbitrap Q Exactive Mass Spectrometer (Thermo Scientific, San Jose, CA, USA) with dual HILIC positive and c18 negative liquid chromatography (Higgins Analytical, Targa, Mountain View, CA, USA, 2.1 x 10 cm) with a formic acid/acetonitrile gradient. The high-resolution mass spectrometer was operated over a scan range of 85 to 1275 mass/charge (*m/z*).(19) Data were extracted and aligned using apLCMS (20) and xMSanalyzer (21) with each feature defined by specific *m/z* value, retention time, and integrated ion intensity.(19) Three technical replicates were performed for each CSF sample and intensity values were median summarized. Identities of targeted metabolites were confirmed by accurate mass, MS/MS and retention time relative to authentic standards.(22)

### Cytokine detection

The commercially available U-PLEX assay by Meso Scale Discovery (MSD) was used for CSF cytokine detection. This assay allows for the evaluation of multiplexed biomarkers by using custom made U-PLEX sandwich antibodies with a SULFO-TAG conjugated antibody and electrochemiluminescence (ECL) detection. Direct quantitation of cytokines was performed using standard curves generated by 4-fold serial dilutions of standard calibrators provided by MSD. Plates were read on the QuickPlex SQ 120 using Methodical Mind^TM^ software and plate data analyzed using Discovery Workbench^TM^ software (MSD).

### Statistics

Statistical comparisons of metabolite intensity values (abundance), metabolite concentrations, and cytokine concentrations were performed in R version 4.2.1. All intensity values and concentrations were log_2_ transformed and compared between groups using linear regression, controlling for age and sex.(23) For untargeted analyses, a false discovery rate (FDR) of q<0.10 was used.(24) Metabolic pathway enrichment analysis was performed using *mummichog*.(25, 26) For correlation analyses, CSF metabolite and cytokine concentrations were normalized using log transformation. Integrative network analyses were performed using xMWAS,(27) which automatically determines the minimum association score threshold required to meet the significance criteria based on the number of samples using Student’s t-test. Only associations with an |association score|≥0.54 significant at p<0.05 were included in the network analysis.

## RESULTS

### Participants

We studied CSF samples from 17 participants with suspected TBM presenting to the NCTLD and 20 asymptomatic persons in the U.S. (**Table 1**). Among the 17 patients with suspected TBM, five (29%) were microbiologically confirmed with a median CSF white blood cell (WBC) count of 209 cells/mm^3^, 94% lymphocytes, a median glucose of 40 mg/dL, and total protein of 99 mg/dL. Five persons with TBM were diagnosed with multi-drug-resistant (MDR)-TBM, two of which were identified through microbiologic testing and three of which were diagnosed clinically based on a poor response to first-line therapy.

**Table 1.**

|  | TB meningitis (n=17) | Controls (n=20) |
| --- | --- | --- |
| Age, median (IQR) | 36 (31-44) | 59 (57-65) |
| Female sex, n (%) | 9 (53) | 10 (50) |
| Microbiologically confirmed TB meningitis, n (%) | 5 (29) | NA |
| CSF total WBC, median (IQR) | 209 (130-286) | NA |
| % Lymphocytes, median (IQR) | 94 (85-96) |  |
| % Neutrophils, median (IQR) | 5 (2-8) |  |
| TB meningitis grade, n (%) |  | NA |
| Grade 1 | 8 (47) |  |
| Grade 2 | 9 (53) |  |

### TB meningitis dramatically alters the CSF metabolome

We detected 6,596 metabolic features in positive ionization mode and 9,427 in negative ionization mode. Using an FDR-adjusted p-value of 0.1 and log2 fold-change threshold of 0.5 (1.414-fold increased or decreased), we found 803 metabolic features were significantly increased in the CSF of TBM patients versus controls (672 in positive ionization mode and 132 in negative mode) while 806 metabolic features were decreased (708 in positive mode and 98 in negative mode; **Figure 1A**). Metabolic pathway analysis revealed vitamin D3, vitamin B3 and multiple amino acid pathways were enriched in the CSF of TBM patients while caffeine metabolism and multiple pathways in carbohydrate metabolism were enriched in the CSF of control participants (**Figure 1B**).

**Figure 1 –.**
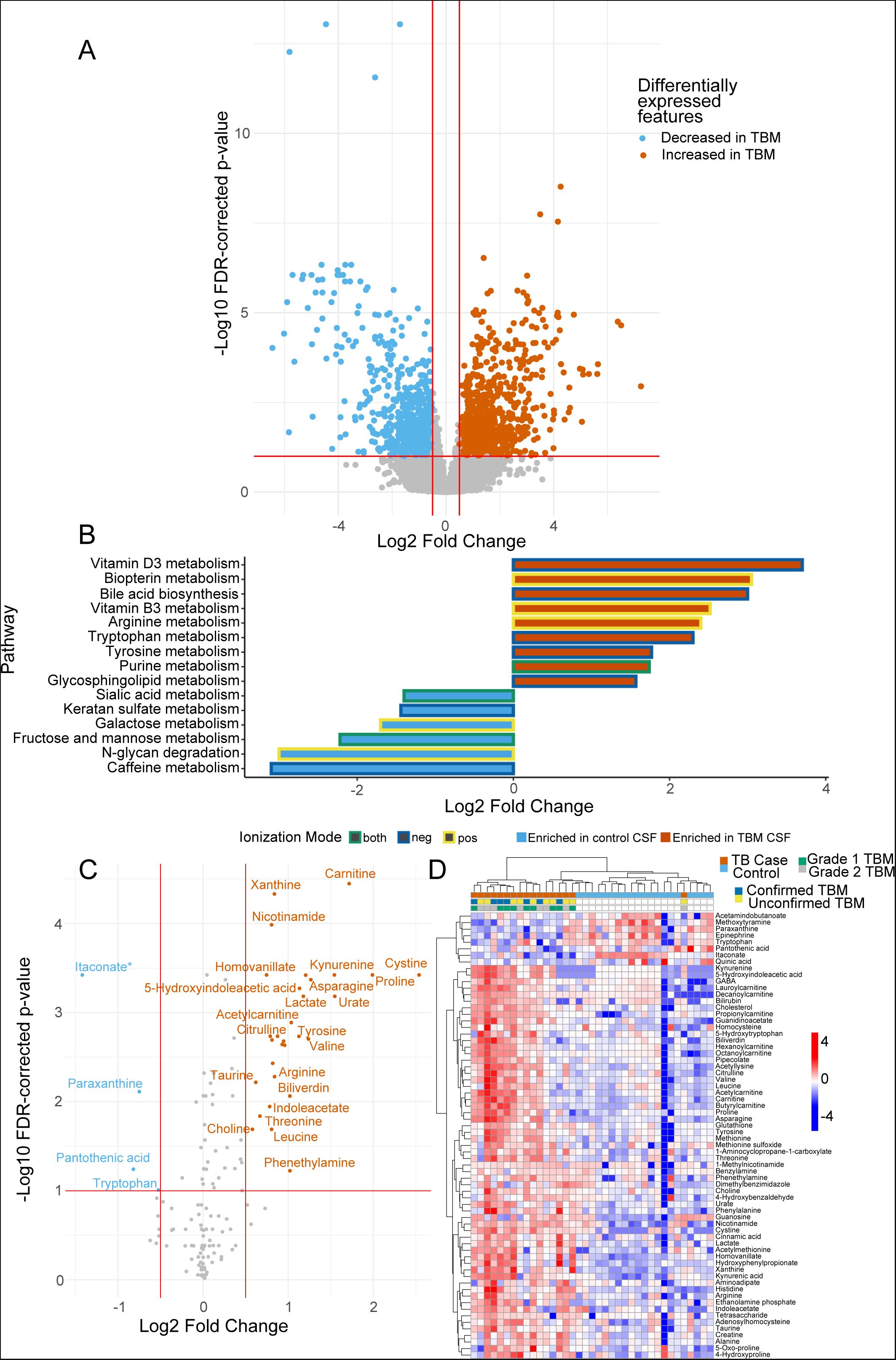
(A) Volcano plot showing m/z features that were differentially abundant in the cerebrospinal fluid of TBM cases versus controls. Those significantly increased in the CSF of TBM cases (log2 fold change ≥ 0.5 and FDR-corrected p-value < 0.1) are in red while those increased in the CSF of control participants are in blue. (B) Metabolic pathway analysis showing metabolic pathways significantly enriched (p<0.05) in TBM CSF (red) and those significantly enriched in control CSF (blue). (C) Volcano plot of targeted metabolites significantly increased in the CSF of TBM cases (red; log2 fold change ≥ 0.5 and FDR-corrected p-value < 0.1) and those increased in the CSF of control participants (blue). *Itaconate is isobaric with citraconate and mesoconate. (D) 2-way hierarchical clustering analysis of targeted metabolites that significantly differed in TBM CSF versus control CSF. Each column is annotated by whether the participant was diagnosed with TBM (red) versus controls (blue), whether TBM was microbiologically confirmed (dark blue) or not (yellow), and whether a TBM case had Grade 1 (grey) versus Grade 2 (green) severity.

To enhance our understanding of the CSF metabolic changes associated with TBM, we performed a targeted analysis of individual metabolites with confirmed chemical identities.(22) Similar to the untargeted analysis, we found the CSF of TBM patients was characterized by severe perturbations in energy metabolism and multiple immunomodulatory metabolites including elevated concentrations of kynurenines, carnitines, amino acids, and lactate and decreases in tryptophan, pantothenic acid, and itaconate (**Figure 1C**). Hierarchical clustering analysis of significant metabolites showed almost all TBM cases formed a single cluster aside from a single, unconfirmed TBM case (**Figure 1D**). Together, these findings suggest a re-ordering of energy metabolism in the CNS of TBM patients, with a metabolic milieu characterized by both glycolysis and lactate accumulation as well as fatty acid activation. Further, we found CSF concentrations of immunoregulatory metabolites tryptophan, kynurenine and itaconate, all of which are known to play an important role in modulating the immune response to pulmonary TB (26, 28–30), were markedly different in TBM cases versus controls.

### Heightened inflammatory cytokines and amino acid metabolism in the CSF identify patients with TB meningitis

We performed a principal component analysis (PCA) of all 33 cytokines detected in the CSF. We found clear separation between the two groups, which was more pronounced in those with confirmed TBM (**Figure 2A**). This was further displayed when visualizing cytokine data using a row normalized heatmap, where TBM cases separated from asymptomatic controls (**Figure 2B**). TBM cases were characterized by marked elevations in most cytokines, and particularly pro-inflammatory cytokines such as IL-17A, TNFα, IFNγ, and IL-1β. Increased expression of effector T cell associated cytokines including IL-2, IFNγ, IL-4, IL-17A and IL-15 (31) was also observed in TBM cases while IL-7, a cytokine associated with homeostasis of naïve and central memory T cells,(32) was decreased.

**Figure 2 –.**
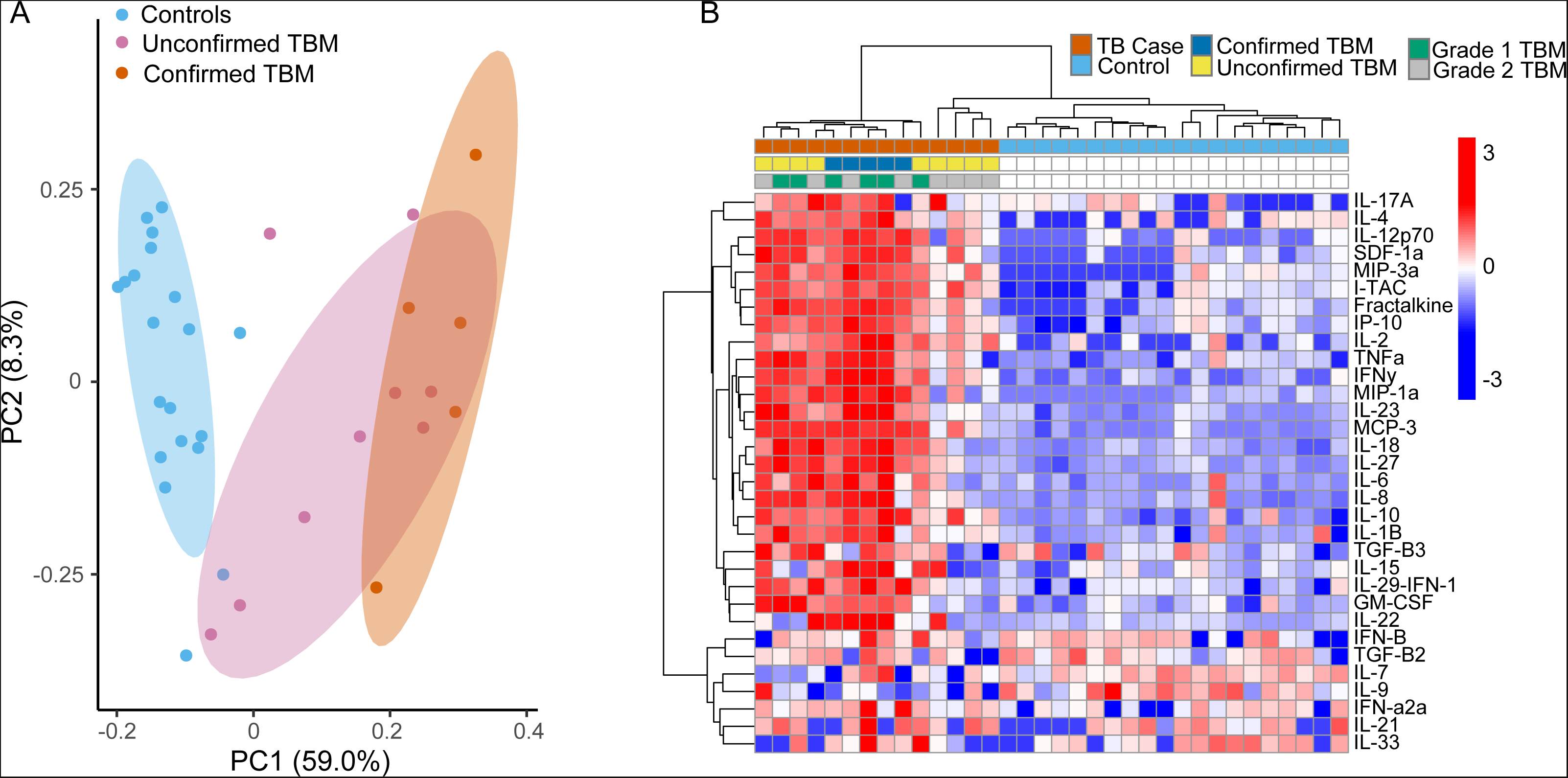
(A) Principal component analysis of all 33 cytokines measured in the CSF showing clear separation between control CSF (blue) and those with microbiologically confirmed TBM (red) while those with unconfirmed TBM (purple) had some overlap with both groups. Principal component 1 (x-axis) captured 59% of the variation in the data while principal component 2 (y-axis) captured 8%. (B) 2-way hierarchical clustering analysis of all measured cytokines annotated by whether the participant was diagnosed with TBM (red) versus controls (blue), whether TBM was microbiologically confirmed (dark blue) or not (yellow), and whether a TBM case had Grade 1 (grey) versus Grade 2 (green) severity.

To determine the relationship between the metabolic and immune response in the CSF, we performed a correlation analysis of metabolite and cytokine concentrations across asymptomatic controls and persons with TBM at diagnosis. This revealed three clusters of metabolites and cytokines respectively (**Figure 3A**). Cluster 1 cytokines consisted of a large group of predominantly pro-inflammatory cytokines upregulated in TBM patients (**Figure 3B**). This cluster was significantly and positively correlated with metabolite cluster 3, primarily consisting of lactate, amino acids, carnitines, and kynurenines (**Figure 3C**). Cluster 1 cytokines were also significantly and negatively correlated with cluster 1 metabolites, which were significantly decreased in TBM cases (**Figure S1A and S1B**). Cluster 2 metabolites and cluster 3 cytokines did not have strong relationships with other soluble mediators, though metabolite cluster 2 was also significantly increased in TBM (**Figure S1C**). Cytokine cluster 2, made up of IL-7 and IL-9, was decreased in TBM cases versus controls (**Figure S2A**) and was weakly correlated with metabolite cluster 1 and weakly inversely correlated with metabolite clusters 2 and 3. While cytokine cluster 3, which consisted of TGFβ2, IFNβ, IFNα2a, IL-21, and IL-33, did not differ in TBM versus controls (**Figure S2B**), it was significantly increased in patients with Grade 2 TBM versus those with Grade 1 TBM (**Figure S2C**), indicating an association with disease severity.

**Figure 3 –.**
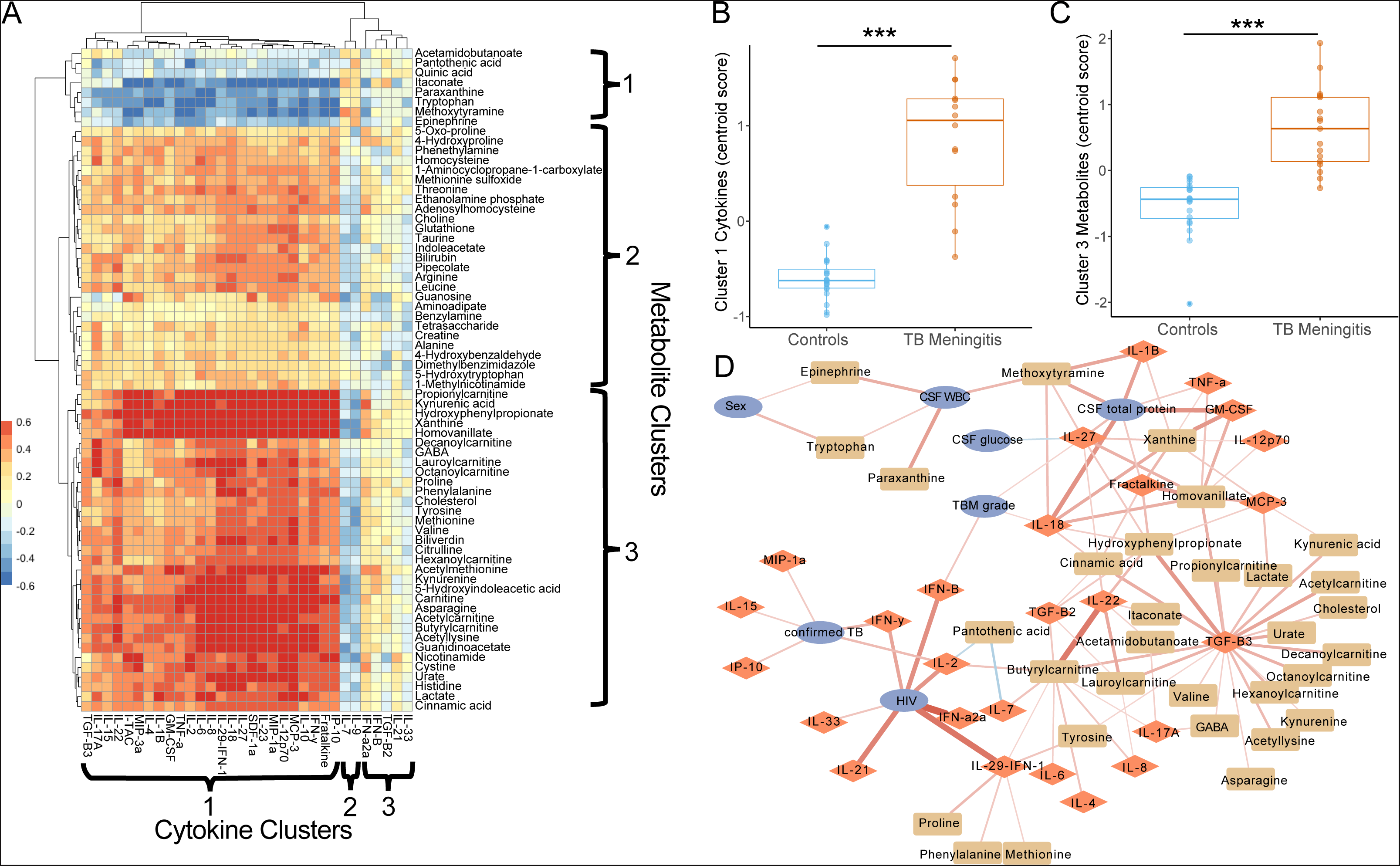
(A) Pearson correlation heatmap of cytokines (x-axis) and differentially abundant metabolites (y-axis) in controls (n=20) and TBM cases at baseline (n=14) with 2-way hierarchical clustering. (B) Comparison of the centroid score of cluster 1 cytokines and (C) cluster 3 metabolites in TBM cases at baseline versus controls. Groups were compared using a Wilcoxon rank sum test (*P ≤ 0.05, **P < 0.01, ***P < 0.001). (D) Unbiased network analysis showing all significant relationships between clinical characteristics of persons diagnosed with TBM and cytokine and metabolite concentrations in CSF at the time of diagnosis. Positive associations are denoted with red edges while negative associations are denoted with blue edges. The intensity and width of the line corresponds to the strength of the association.

To determine the network structure between clinical variables and CSF metabolite and cytokine concentrations in TBM, we used a platform-independent framework for integrative network analysis called xMWAS.(27) Using this approach we found TGFβ3 played a central role in the activity network, with significant associations with multiple metabolites including kynurenines and carnitines (**Figure 3D**). While there was only one participant with TBM that was HIV positive, HIV was significantly associated with type I interferons and IFNγ. Confirmed TBM was associated with IL-2, IFNγ, MIP-1α, IL-15, and IP-10 while TBM grade was associated with IFNβ, IL-18, and IL-27. Tryptophan and epinephrine were associated with increased CSF WBC count.

### Inflammatory immunometabolic changes in TBM persists for months despite appropriate treatment

We next examined the trajectory of CSF metabolite and cytokine concentrations after initiation of TBM treatment. Repeat CSF samples were examined 7, 14, 28 and 56 days after treatment initiation. Of the 17 TBM patients enrolled, 3 patients were eventually deemed “non-responders” due to suspected or proven drug resistance, while 14 patients improved clinically on anti-TB therapy. Treatment responders had significant decreases in cluster 1 cytokines at 14, 28, and 56 days after treatment initiation (**Figure 4A**). However, after 56 days of appropriate treatment, CSF concentrations of these cytokines remained significantly higher than in asymptomatic controls. After 7 and 14 days of treatment, the three treatment non-responders had higher concentrations of cluster 1 cytokines versus responders, though the difference was not statistically significant (**Figure 4B**). Cluster 2 cytokines were lower in TBM patients versus controls and did not increase with treatment (**Figure S3A**) while cluster 3 cytokines were similar to controls at baseline and throughout treatment (**Figure S3B**).

**Figure 4 –.**
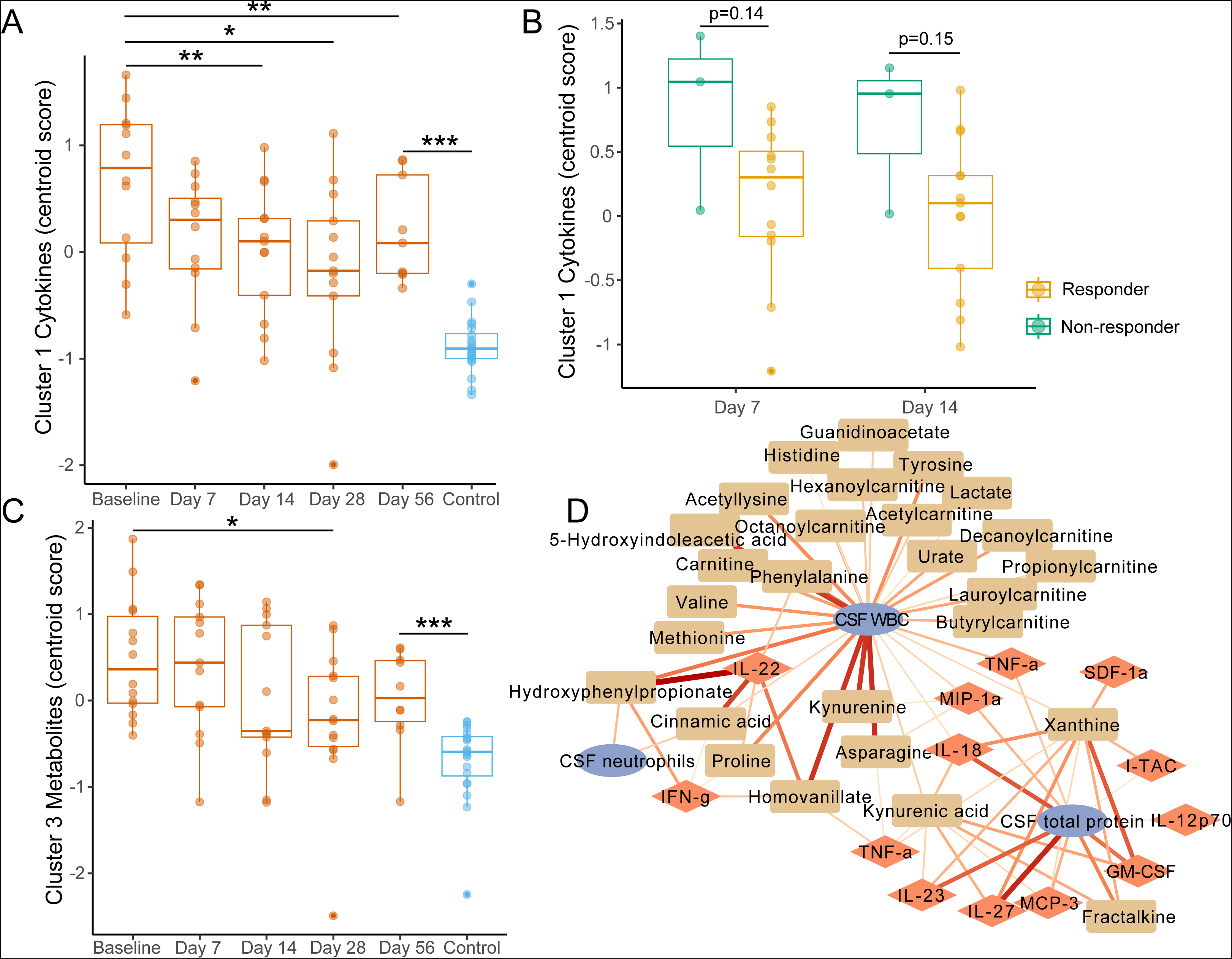
(A) Boxplots showing changes in cluster 1 cytokines after 7, 14, 28, and 56 days of treatment in persons with TBM who clinically responded to therapy. (B) after 7 and 14 days, treatment non-responders had elevated concentrations of cluster 1 cytokines versus responders. (C) Cluster 3 metabolites also significantly declined in the CSF in treatment responders. Cross sectional measures were compared with a Wilcoxon rank sum test while repeat measures used a Wilcoxon signed rank test: * p<0.05, ** p<0.01, *** p<0.001. (D) Unbiased network analysis showing all significant relationships between clinical characteristics of persons diagnosed with TBM and cytokine and metabolite concentrations in CSF during treatment. Positive associations are denoted with red edges while negative associations are denoted with blue edges. The intensity and width of the line corresponds to the strength of the association.

In evaluating the metabolome over the course of TBM treatment, we fount cluster 1 metabolites did not increase with TBM treatment and remained significantly lower than controls after 56 days (**Figure S4A**). Cluster 1 metabolites were lower in non-responders versus responders after 7 days of treatment, though the difference was not statistically significant (**Figure S4B**). Conversely, cluster 2 metabolites were significantly lower after 28 days of treatment (**Figure 4C**) and cluster 3 metabolites were significantly lower after 28 and 56 days of treatment (**Figure S4C**). However, both remained significantly elevated after 56 days of treatment relative to controls. Network analysis of metabolites and cytokines during TBM treatment revealed that metabolites had the strongest association with CSF WBC count during treatment, including lactate, kynurenine, amino acids, and carnitines (**Figure 4D**). Concentrations of MIP-1α, IL-18 and TNFα were also significantly associated with CSF WBC count.

## DISCUSSION

In this study, we show that multiple metabolic signaling networks are closely associated with the CNS inflammatory response in patients with TBM and that even after initiating TBM treatment, these immunometabolic changes last for an extended duration. Over the course of treatment, it was metabolites, including kynurenine and multiple carnitines, that were most closely associated with elevated WBCs in the CSF rather than cytokines. These findings indicate that metabolic shifts in the CSF of TBM patients are an integral part of the host inflammatory response. Despite recent advances in the treatment of TB disease,(33–35) morbidity and mortality in persons with TBM remains unacceptably high.(1, 3) There is increasing evidence that a dysregulated immune response in the CNS is a primary driver of poor patient outcomes.(3, 36) Our findings strongly support the hypothesis that chronic inflammation and immune dysfunction in the CNS persist despite effective TB treatment and that this may be the root cause of neurological complications, morbidity, and mortality. Studying the molecular networks that modulate pathologic inflammation in TBM is therefore essential to discover strategies to ameliorate inflammatory immunometabolomic changes in the CNS.

The simultaneous measurement of 33 cytokines and chemokines in the CSF is, to our knowledge, the most comprehensive assessment of CSF soluble immune mediators in TBM. Similar to previous studies measuring a smaller number of cytokines, we observed a significant shift in the CSF cytokine profile when we compared individuals with TBM to asymptomatic adults.(8, 9) Among the cytokines that were upregulated in the CSF of TBM patients were key modulators of inflammatory T cell and innate immune responses including IFNγ, TNFα, IL-6, GM-CSF, IL-18, and IL-1β. We also found that TBM was associated with elevation in multiple anti-inflammatory cytokines including IL-10 and TGF-β3. This suggests that both inflammatory pathways triggered by infection and their counter-regulatory response networks are increased in TBM. In our integrative analysis of TBM cases at the time of diagnosis, chemokines IP-10 and MIP-1α as well as cytokines IFNγ, IL-15, and IL-2 were significantly associated with confirmed TBM, indicating these soluble mediators are particularly active in TBM cases with higher bacterial burden. The network analysis revealed the cytokines IFNβ, IL-27, and IL-18 were significantly associated with higher TBM grade, suggesting a potential role in mediating immunopathology in severe TBM.

We found the concentration of multiple immunomodulatory metabolites in the CNS differentiated persons with TBM from asymptomatic controls. Notably, the metabolites itaconate and tryptophan were two of the few metabolites significantly decreased in the CSF of TBM patients versus controls in the targeted analysis. Itaconate concentrations were significantly and inversely correlated with concentrations of most inflammatory cytokines and chemokines including IL-1β, IL-18, TNFα, IFNγ, MIP-1α, and MIP-3α. Itaconate is derived from TCA cycle intermediate cis-aconitate and has been shown to modulate inflammatory signaling in macrophages through inhibition of succinate dehydrogenase (37), induction of Nrf2 (38), and inhibition of inflammasome activation.(39) We have previously shown that itaconate is decreased in plasma of humans with advanced pulmonary TB disease,(28) and mice lacking the enzyme that forms itaconate experience pathologic pulmonary inflammation and rapid mortality.(30) Itaconate has also been shown to inhibit IL-1β-mediated inflammation in astrocytes(40) and microglia,(41) the latter of which is a primary source of inflammasomes in the CNS.(42) This suggests low CSF itaconate may play a role in an overexuberant CNS inflammatory response. Similar to a previous TBM metabolomics study, we also found that persons with TBM had decreased CSF concentrations of tryptophan(13) and increased concentrations of its immediate metabolite, kynurenine. Induction of tryptophan catabolism via the enzyme indoleamine-2,3-dioxygenase-1 (IDO1) is a hallmark of TB disease regardless of the affected organ system.(26) Kynurenine and its metabolites are ligands of the aryl hydrocarbon receptor (AHR), which promotes differentiation of regulatory T cells (Tregs) at the expense of Th17 cells.(43–45). This has the effect of limiting T cell effector function and inflammation. This effect is likely protective in TBM, where decreased T cell activation would be expected to reduce immunopathology. Indeed, increased tryptophan catabolism in the CNS of persons with TBM has been associated with increased survival.(13, 14) We observe significantly elevated Th1 (IL-2, IFNγ), Th2 (IL-4) and Th17 (IL-17, IL-6, TNFα) effector cytokines in the CSF of persons with TBM,(31) and these cytokines were correlated with CSF concentrations of kynurenine and kynurenic acid. This indicates tryptophan catabolism represents a counter-regulatory process to limit T-cell-mediated inflammation.(46)

We also found metabolic changes in the CSF of TBM patients that were consistent with broad shifts in energy metabolism. Persons with TBM had significant increases in multiple carnitines as well as lactate, suggesting simultaneous activation of beta-oxidation of fatty acids and glycolysis. In the integrative analysis, these metabolic networks were all significantly associated with TGFβ signaling. Such metabolic changes may therefore stimulate TGFβ secretion or occur as a result of TGFβ upregulation. During treatment lactate, kynurenine, and carnitines, along with IL-18, MIP-1α, and TNFα were significantly associated with CSF WBC count. Elevated expression of chemokines promotes the migration of myeloid and lymphoid cells into the CSF where they are then activated for inflammatory cellular responses by cytokines including IL-1β, IL-6, IL-18, TNFα, and IFNγ. Our data provides the framework for a potential mechanism of dysregulated immune cell infiltration and activation in the CSF that is driven by metabolic changes during TBM treatment.

The immunometabolic shifts seen in the CSF of TBM patients showed only minor signs of resolution after two months of effective treatment. In cluster 1 metabolites, which included tryptophan and itaconate, there was no increase over the first 2 months of treatment and these metabolites remained significantly lower than controls. While there was a significant decrease in metabolite cluster 3 with treatment, which included carnitines, kynurenines, and lactate, these still remained significantly elevated relative to controls. Similarly, while proinflammatory cytokines and chemokines (cluster 1) decrease with effective TBM treatment, these cytokines remained significantly elevated relative to controls after two months of treatment. This prolonged state of abnormal immunometabolic signaling follows with clinical observations that mortality in TBM patients remains elevated months to years after treatment initiation.(47) It also mimics what is seen for many chronic inflammatory conditions where normal immune homeostasis is permanently altered even after removal of the underlying cause of the disease.(48–50) This strongly supports consideration of TBM as a chronic inflammatory condition which may require longer-term immunomodulatory treatment.

This study is subject to several limitations. Cross-sectional comparison of CSF metabolite and cytokine concentrations in TBM patients were made to a control group of asymptomatic individuals. This makes it more difficult to distinguish which molecular signatures are specific to TBM versus meningitis due to other causes. Additionally, the relatively small sample size of TBM cases from a single geographic region may limit the generalizability of the study. Though we demonstrate a strong association between CSF concentrations of multiple metabolites and cytokines/chemokines, the observational nature of the study precludes us from establishing a causal relationship. In future studies it will be important to combine systemic measurement of soluble immune modulators in the CSF with transcriptional profiling of immune cells in the CNS to better understand the immune cell phenotypes and signaling pathways associated with pathologic metabolic signaling in TBM. Additionally, mechanistic validation of the link between key metabolites and inflammatory mediators can be assessed using *ex vivo* or *in vitro* stimulation systems.

Overall, the data presented clearly demonstrate that soluble factors (cytokines and metabolites) are critical features of the host immune environment which distinguish the presence and severity of TBM. Our results provide insight into the metabolic drivers of CSF inflammation in TBM, which are an integral part of inflammatory signaling networks. Given the role of immunopathology in mediating adverse host outcomes in TBM, this will be a critical area for further research. As these findings are expanded, it can be envisioned that signatures of these host molecules could have utility as biomarkers of TBM, disease severity, and response to TB treatment.

## Data Availability

All data produced in the present study are available upon reasonable request to the authors.

## Acknowledgments

The authors thank the physicians, nurses, and staff at the NCTLD in Tbilisi, Georgia, who provided care for the patients with TBM included in this study. Additionally, the authors are thankful for study participants with tuberculosis meningitis who were willing to participate in the study and help contribute meaningful data that may help future patients with the same illness.

## Conflict of Interest

The authors declare that the research was conducted in the absence of any commercial or financial relationships that could be construed as a potential conflict of interest.

## Author Contributions

All authors agree to be accountable for the content of the work. JT and JC led the data analysis, integration, and interpretation, as well as manuscript writing. JT and AS led the cytokine analysis and JC led the metabolomics analysis. TB and SS provided the care for all patients with TBM at the National Center for Tuberculosis and Lung Diseases and helped in data collection and reviewed the manuscript drafts. AGCS assisted with data collection and preparation and reviewed the manuscript drafts. MK and RRK led and designed the parent clinical study, led project implementation, and assisted with data interpretation and reviewing the manuscript.

## Funding

This work was supported by grants from the National Institutes of Health (NIH) and National Institute of Allergy and Infectious Diseases [R03AI139871, K23AI103044, K23AI144040, P30AI168386, P30AI050409]; NIH Fogarty International Center [D43TW007124]; and NIH National Center for Advancing Translational Science [UL1TR002378], Bethesda, MD, USA.

## Figure Legends

**Supplemental Figure 1 –** (A) Cluster 1 metabolites including (B) tryptophan were significantly decreased in the CSF of persons diagnosed with TBM versus asymptomatic controls. (C) Cluster 2 metabolites were significantly increased in TBM versus asymptomatic controls. Groups were compared using a Wilcoxon rank sum test (*P ≤ 0.05, **P < 0.01, ***P < 0.001).

**Supplemental Figure 2 –** (A) Cluster 2 cytokines (IL-7 and IL-9) were significantly lower in the CSF of TBM patients versus asymptomatic controls while (B) Cluster 3 cytokines did not differ between groups. (C) Cluster 3 cytokines were significantly increased in those with Grade 2 TBM versus those with Grade 1 TBM. Groups were compared using a Wilcoxon rank sum test (*P ≤ 0.05, **P < 0.01, ***P < 0.001).

**Supplemental Figure 3 –** (A) Cluster 2 cytokines did not increase in the CSF in treatment responders after 7, 14, 28, and 56 days of treatment and remained significantly lower than asymptomatic controls. (B) Cluster 3 cytokines did not change with TBM treatment and remained similar to concentrations in the CSF of asymptomatic controls. Cross sectional measures were compared with a Wilcoxon rank sum test while repeat measures used a Wilcoxon signed rank test: * p<0.05, ** p<0.01, *** p<0.001.

**Supplemental Figure 4 –** (A) Cluster 1 metabolites did not increase in the CSF in treatment responders after 7, 14, 28, and 56 days of treatment and remained significantly lower than asymptomatic controls. (B) Cluster 1 metabolites were lower in treatment non-responders after 7 days of treatment versus treatment responders. (C) Cluster 2 metabolites significantly decreased over the course of TBM treatment but remained significantly increased versus asymptomatic controls. Cross sectional measures were compared with a Wilcoxon rank sum test while repeat measures used a Wilcoxon signed rank test: * p<0.05, ** p<0.01, *** p<0.001.

## Notes

### Competing Interest Statement

The authors have declared no competing interest.

### Funding Statement

This work was supported by grants from the National Institutes of Health (NIH); National Institute of Allergy and Infectious Diseases [R03AI139871, K23AI103044, K23AI144040, P30AI168386, P30AI050409]; NIH Fogarty International Center [D43TW007124]; and NIH National Center for Advancing Translational Science [UL1TR002378], Bethesda, MD, USA.

### Author Declarations

The institutional review boards of Emory University and the National Center for Tuberculosis and Lung Disease gave ethical approval for this work.

