## Supplementary figures and images for "Combined cerebrospinal fluid metabolomic and cytokine profiling in tuberculosis meningitis reveals robust and prolonged changes in immunometabolic networks"

### Supplemental Figures

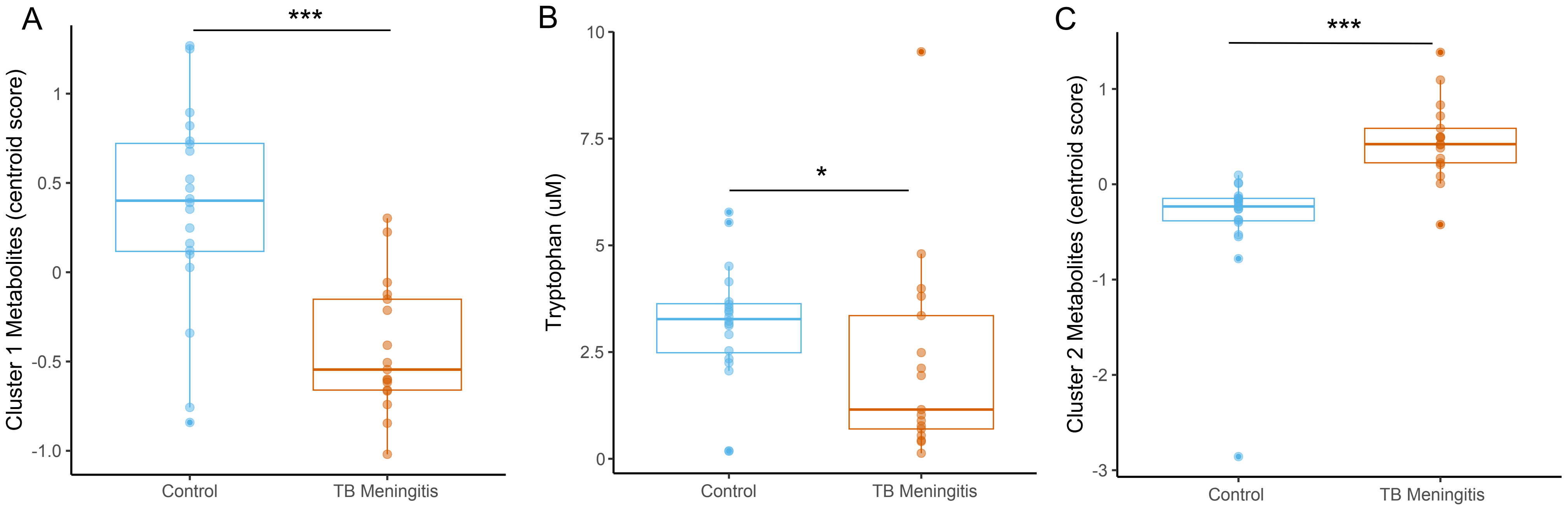

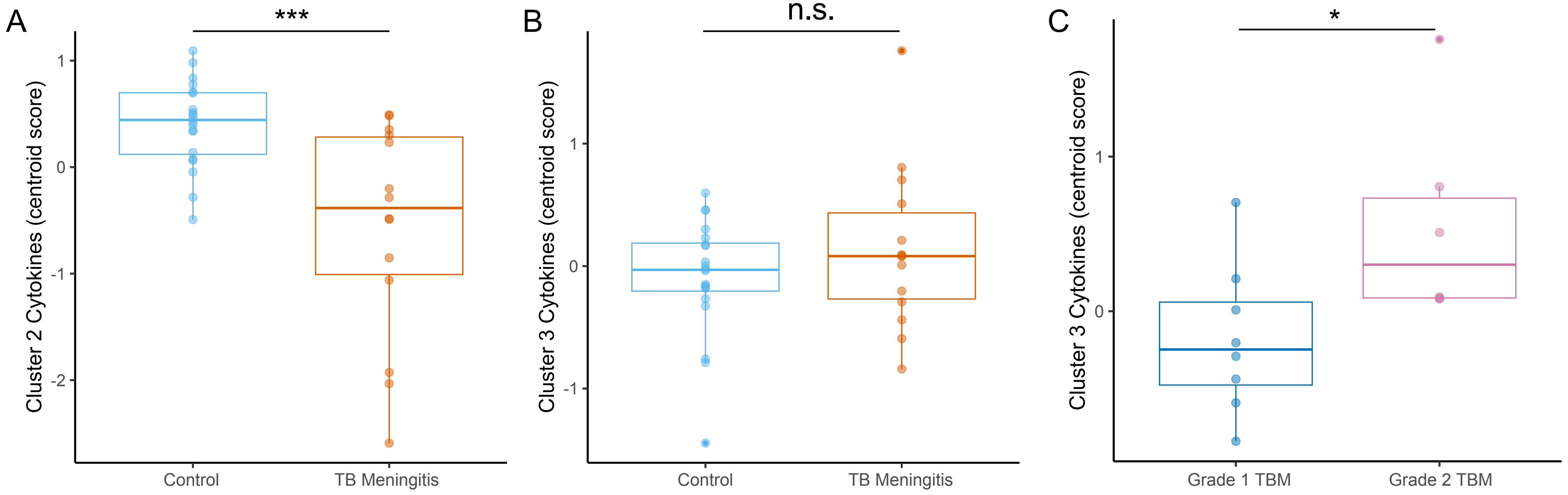

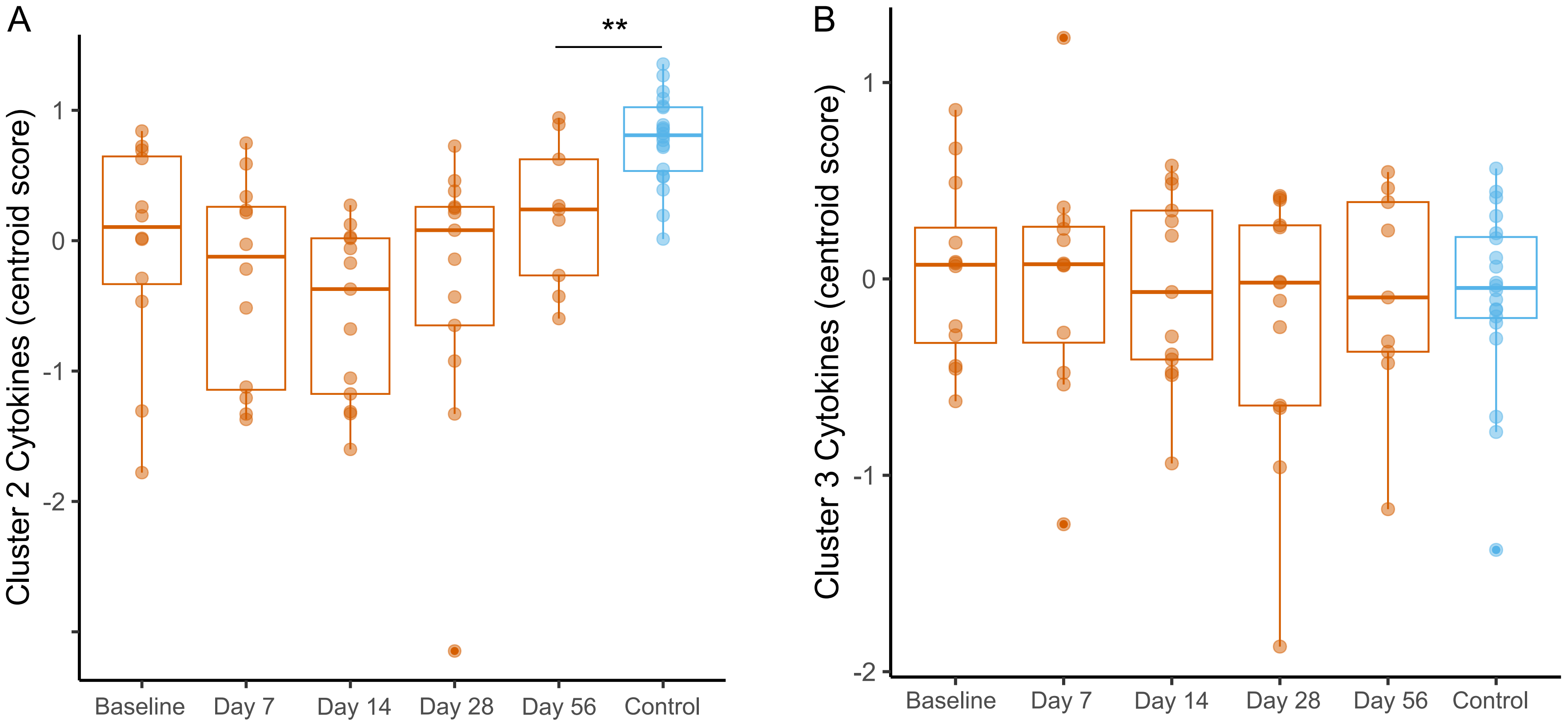

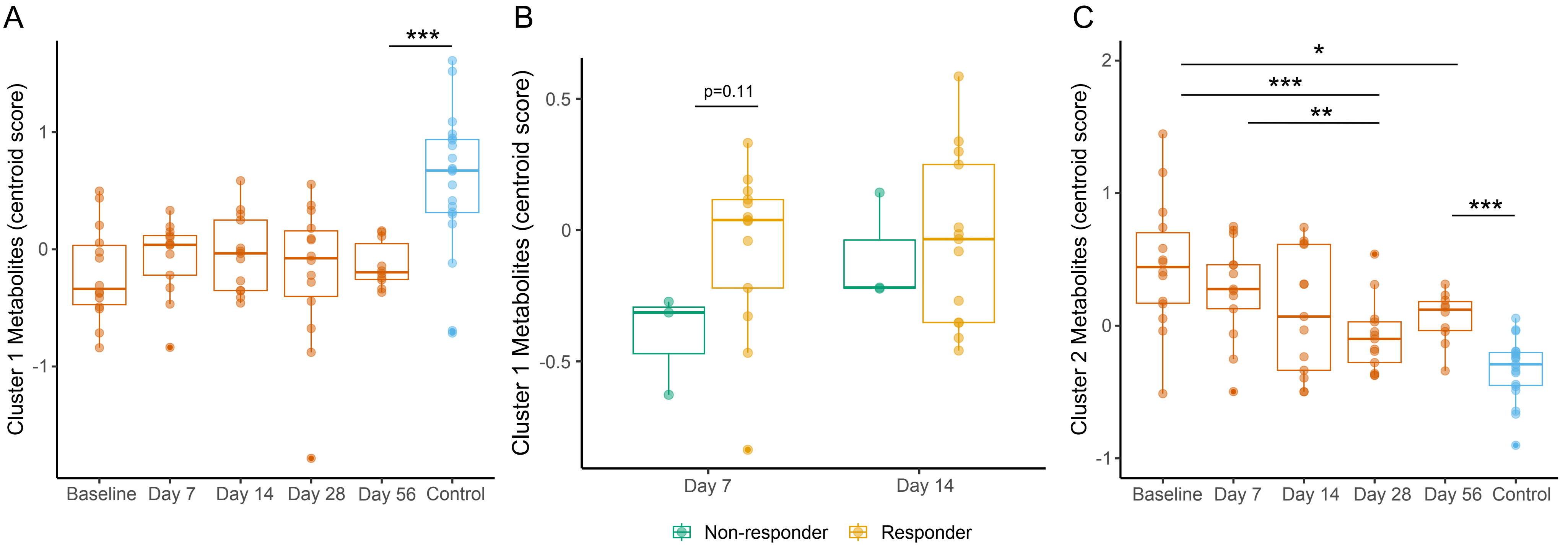
